# Automated Tumor Segmentation and Brain Tissue Extraction from Multiparametric MRI of Pediatric Brain Tumors: A Multi-Institutional Study

**DOI:** 10.1101/2023.01.02.22284037

**Authors:** Anahita Fathi Kazerooni, Sherjeel Arif, Rachel Madhogarhia, Nastaran Khalili, Debanjan Haldar, Sina Bagheri, Ariana M. Familiar, Hannah Anderson, Shuvanjan Haldar, Wenxin Tu, Meen Chul Kim, Karthik Viswanathan, Sabine Muller, Michael Prados, Cassie Kline, Lorenna Vidal, Mariam Aboian, Phillip B. Storm, Adam C. Resnick, Jeffrey B. Ware, Arastoo Vossough, Christos Davatzikos, Ali Nabavizadeh

## Abstract

**Background:** Brain tumors are the most common solid tumors and the leading cause of cancer-related death among all childhood cancers. Tumor segmentation is essential in surgical and treatment planning, and response assessment and monitoring. However, manual segmentation is time-consuming and has high interoperator variability. We present a multi-institutional deep learning-based method for automated brain extraction and segmentation of pediatric brain tumors based on multi-parametric MRI scans.

**Methods:** Multi-parametric scans (T1w, T1w-CE, T2, and T2-FLAIR) of 244 pediatric patients (n=215 internal and n=29 external cohorts) with de novo brain tumors, including a variety of tumor subtypes, were preprocessed and manually segmented to identify the brain tissue and tumor subregions into four tumor subregions, i.e., enhancing tumor (ET), non-enhancing tumor (NET), cystic components (CC), and peritumoral edema (ED). The internal cohort was split into training (n=151), validation (n=43), and withheld internal test (n=21) subsets. DeepMedic, a three-dimensional convolutional neural network, was trained and the model parameters were tuned. Finally, the network was evaluated on the withheld internal and external test cohorts.

**Results:** Dice similarity score (median±SD) was 0.91±0.10/0.88±0.16 for the whole tumor, 0.73±0.27/0.84±0.29 for ET, 0.79±19/0.74±0.27 for union of all non-enhancing components (i.e., NET, CC, ED), and 0.98±0.02 for brain tissue in both internal/external test sets.

**Conclusions:** Our proposed automated brain extraction and tumor subregion segmentation models demonstrated accurate performance on segmentation of the brain tissue and whole tumor regions in pediatric brain tumors and can facilitate detection of abnormal regions for further clinical measurements.

**Key Points:**

- We proposed automated tumor segmentation and brain extraction on pediatric MRI.
- The volumetric measurements using our models agree with ground truth segmentations.

**Importance of the Study:** The current response assessment in pediatric brain tumors (PBTs) is currently based on bidirectional or 2D measurements, which underestimate the size of non-spherical and complex PBTs in children compared to volumetric or 3D methods. There is a need for development of automated methods to reduce manual burden and intra- and inter-rater variability to segment tumor subregions and assess volumetric changes. Most currently available automated segmentation tools are developed on adult brain tumors, and therefore, do not generalize well to PBTs that have different radiological appearances. To address this, we propose a deep learning (DL) auto-segmentation method that shows promising results in PBTs, collected from a publicly available large-scale imaging dataset (Children’s Brain Tumor Network; CBTN) that comprises multi-parametric MRI scans of multiple PBT types acquired across multiple institutions on different scanners and protocols. As a complementary to tumor segmentation, we propose an automated DL model for brain tissue extraction.

## Introduction

Pediatric brain tumor (PBT), the most frequent solid tumor and the leading cause of cancer-related death in children, encompasses numerous distinct histologic types ^1^. With the emergence of novel therapies for PBTs, accurate and reproducible tumor assessment metrics that provide early evidence of response or relapse, and therefore, the efficacy of the administered treatment, are essential. The response assessment in pediatric neuro-oncology (RAPNO) working group has developed consensus recommendations for response assessment using measurements in two perpendicular planes (bidirectional or 2D) in several PBT types to be applied in clinical trials and routine neuro-oncology practice ^2–4^. These criteria are built on the assumption that tumor growth is accompanied with an increase in the major axis of the tumor and therefore, bidirectional measurement can serve as a surrogate for tumor volume ^5^.

This assumption may not be valid in many cases when the tumor undergoes irregular growth as a result of treatment-induced effects. Studies on adult brain tumors, have shown that three-dimensional (3D) or volumetric measurements better predict tumor burden and response, compared to the 2D measurements recommended by response assessment in neuro-oncology (RANO) working group ^6,7^. As PBTs are generally non-spherical and often have complex, mixed solid and cystic components ^4^, their assessment requires manual segmentation to quantify and differentiate tumor subregions. However, manual segmentation is not practical in clinical applications, as it is time consuming and prone to intra- and inter-rater variability ^8^.

The current state-of-the-art approach for automated brain tumor segmentation is deep learning (DL); however, most available auto-segmentation tools have been trained and made available for use in adult cancers only ^9–14^. Such models do not generalize well to PBTs ^15^ due to the different radiological appearance of tumors compared to adult brain tumors, and the anatomical differences as a result of the developing brain in children ^16^. A few studies have proposed different DL solutions to the PBT segmentation problem, with whole tumor Dice scores ranging between 0.68-0.76 ^17–20^. These approaches show lower performance than the models proposed for adult brain tumor segmentation, are often only designed for a particular histology, only segment the whole tumor without subregions, or segment the tumors based on one or two MRI sequences. Thus, there remains an unmet need for a broader auto-segmentation method that is specifically designed for PBTs with higher accuracy. It would ideally work for a variety of histologies and tumor locations, segment tumor subregions, and use information from multiple MRI sequences important for distinction of tumor subregions.

To address this need, in the present work, we developed a scalable and fully automated segmentation method using deep learning (DL) based on 3D convolutional neural networks (CNNs) for whole tumor (WT) delineation and differentiation of tumorous subregions, i.e., enhancing tumor and non-enhancing core (including non-enhancing tumor, cystic components, and peritumoral edema) on multiparametric MRI (mpMRI) scans. Furthermore, as a key part of image processing and automated tumor segmentation pipelines, we trained an automated DL-based brain tissue extraction model (“skull-stripping”) using mpMRI scans in presence of tumor. This essential step is intended to overcome the shortcomings of the currently available tools in accounting for structural variability in the brains of developing children, as well as anatomical distortions caused by tumors not addressed by skull-stripping methods based on healthy brain atlases.

## Methods

### Patient Cohort

Our retrospective study was compliant with the HIPAA and obtained approval from the IRB of the Children’s Hospital of Philadelphia (CHOP) through Children’s Brain Tumor Network (CBTN) Protocol. MRI exams were collected from the subjects enrolled onto the CBTN, which is a biorepository (cbtn.org ^21^) that allows for the collection of specimens, longitudinal clinical and imaging data, and sharing of de-identified samples and data for future research. From this dataset, multi-parametric MRI scans of n = 273 patients with histologically confirmed pediatric brain tumors were selected. Patients were excluded if they lacked any of the four standard scans (pre-contrast T1w, post-contrast T1w (T1w-CE), T2w, and T2-FLAIR), their scans had significant imaging artifacts, or the scans were not acquired prior to surgical resection or treatment. However, patients were included when the only available scan sessions were after a biopsy procedure or external ventricular drain (EVD) placement without any other interventions, and without significant malformation of brain skull and/or parenchyma. A final number of 244 subjects (215 from internal site, and 29 from external sites) were included in this study. Table S1 (Supplementary Materials) provides a detailed description of patients’ characteristics.

### Data Pre-Processing, Expert Brain Tissue Extraction, and Tumor Subregion Segmentation

For each patient, T1w, T2w, and T2-FLAIR images were co-registered with their corresponding T1w-CE scan and resampled to an isotropic resolution of 1 mm^3^ based on anatomical SRI24 atlas ^22^ using a previously described greedy algorithm ^23^. Preliminary brain masks and tumor segmentations were generated using DL approaches based on DeepMedic architecture trained on MRI scans of adult GBMs for skull stripping and tumor segmentation, provided in CaPTk ^10,11^. The brain masks were then manually revised based on T1w and T2w scans by medical students experienced in neuroimaging in ITK-SNAP ^24^. In a few pre-segmentation training and consensus sessions, three expert neuroradiologists (A.N. with 10, A.V. with 16, and J.B.W. with 6 years of experience) reviewed images of a subset of patients to provide plans and guidelines for segmentations of four tumor subregions, including enhancing tumor (ET), non-enhancing tumor (NET), cystic component (CC), and edema (ED). As in all tumor segmentation tasks, differentiation of enhancing tumor, non-enhancing tumor, and edema can be challenging. Therefore, difficult cases were reviewed and finalized by consensus among all three neuroradiologists.

Starting from preliminary tumor segmentations, manual revisions were performed using all four MRI scans by several researchers, trained for segmentation of pediatric brain tumors by expert neuroradiologists, to delineate the four tumor subregions. Finally, the neuroradiologists reviewed, manually edited as needed, and finalized the tumor segmentations. These manual segmentations were used as the ground truth in subsequent model training, validation, and performance analysis. Figure 1 illustrates an example of expert segmentation of brain tissue and tumor subregions in a sample patient with the segmentation labels used in this study.

**Figure 1.**
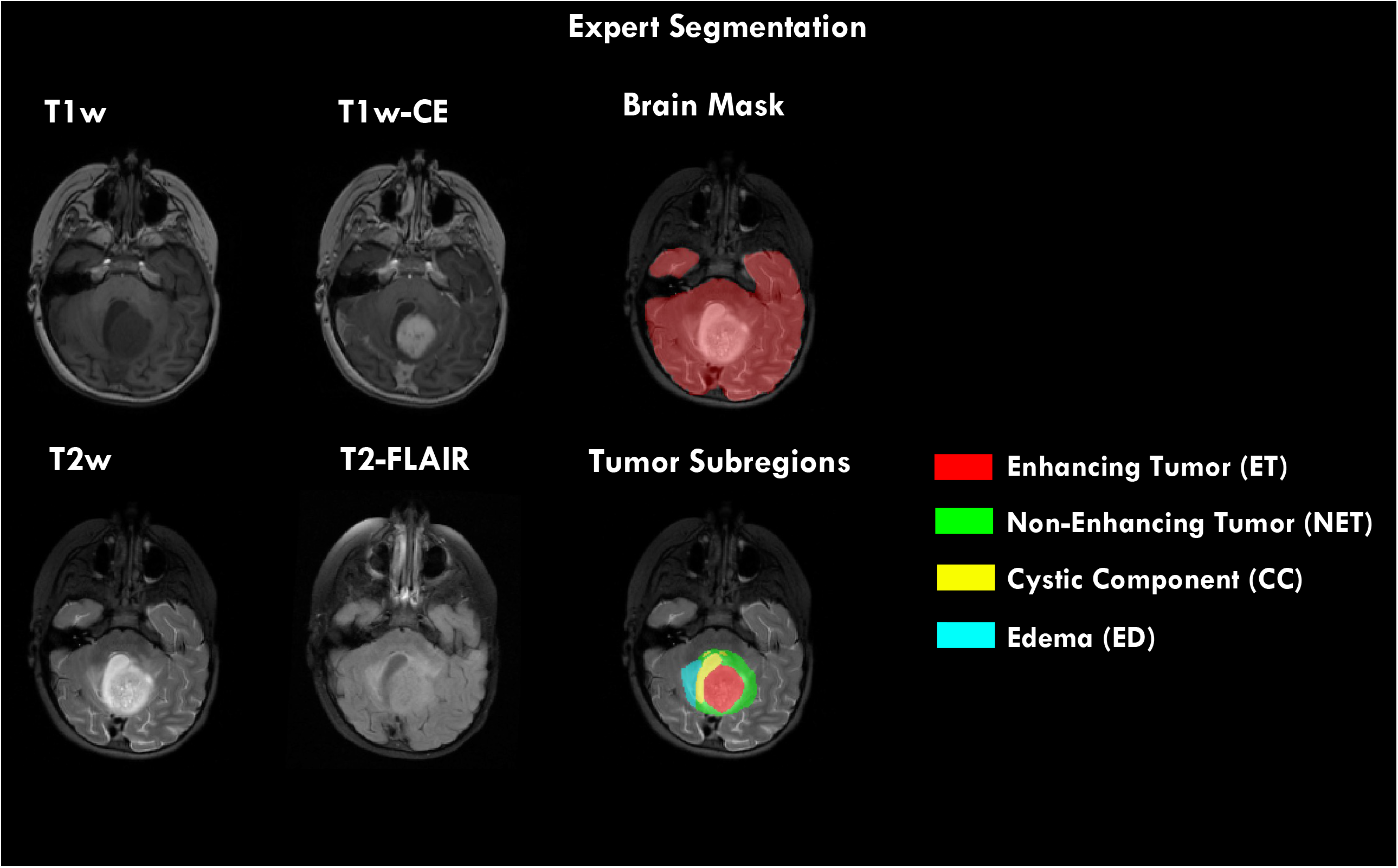
Expert segmentation on a sample patient illustrating how brain tissue and tumor subregions were segmented.

### Model Training

DeepMedic^17,18^ is a 3D convolutional neural network (CNN) that processes images at multiple scales simultaneously. One pathway processes images at their normal resolution, and two additional parallel pathways process images at subsampled resolutions, before the pathways are concatenated. DeepMedic (v0.7.1, see https://github.com/deepmedic/deepmedic) was trained after the patient cohort from the internal site (n = 215) was randomly split into 70%-20%-10% training-validation-test sets. The input to the tumor subregion segmentation consisted of multiparametric MRI scans (T1w, T1w-CE, T2, and T2-FLAIR) along with the brain tissue mask. Images were normalized to zero-mean, unit-variance prior to input into the CNNs. The optimum value for learning rate was 0.001, the number of epochs, 35, and the batch size, 10.

### Performance Analysis

Model performance was assessed by comparing predicted segmentations to their corresponding expert segmentations. Sørensen-Dice score, sensitivity, and 95% Hausdorff distance metrics were calculated for brain tissue (for the skull-stripping model), whole tumor (WT; defined as the union of all four subregions), enhancing tumor (ET), and non-enhancing core (NEC; the combination of all non-enhancing regions, i.e., non-enhancing tumor, cystic components, and edema). NEC label was generated to validate the performance of the model in distinguishing non-enhancing from enhancing areas more globally, particularly as edema and cystic components are infrequent in many subjects. To further evaluate the agreement between the predicted and expert tumor segmentations, a few semantic radiomic features were calculated, some of which are included among VASARI (Visually AcceSAble Rembrandt Images)^20^ features, including proportion of the whole tumor volume that is enhancing tumor, non-enhancing tumor, cystic components, or edema. Pearson’s correlation coefficient (significance level p < 0.05) was calculated between the VASARI features obtained via predicted or expert segmentations.

## Results

In Figure 2, two examples of predicted brain tissue and tumor subregion segmentation as compared to the expert segmentations (used as the ground truth (GT)) are shown. The following sub-sections present the detailed evaluation of skull-stripping and tumor subregion segmentation models.

**Figure 2.**
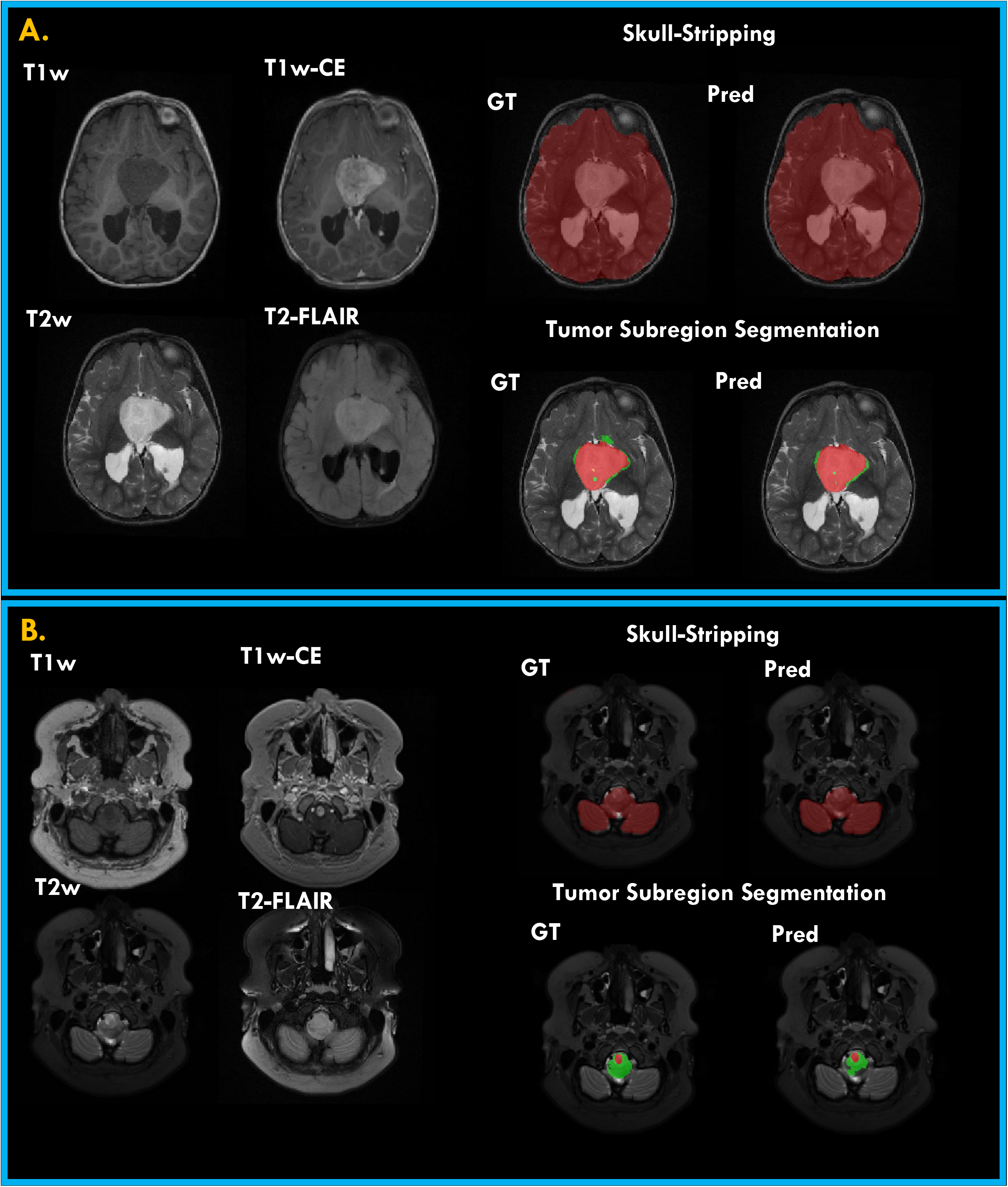
Examples of predicted (Pred) and expert or ground truth (GT) segmentations for skull-stripping and tumor subregions in two patients. (A) a female in age range of 0-4 years with a hypothalamic low-grade astrocytoma; (B) a female with low-grade astrocytoma in medulla in age range of 11-18 years.

### Skull-Stripping

The predicted brain tissue extraction results were compared to those obtained through manual segmentation (as the ground truth), by calculating the performance metrics, i.e., Dice score, sensitivity, and 95% Hausdorff distance in the validation, withheld internal test, and external test sets (reported in Table 1). The proposed algorithm showed high median Dice score of 0.98 ± 0.01 in the validation set, with high reproducibility and generalizability in the withheld internal test (Dice = 0.98 ± 0.02) and external test (Dice = 0.98 ± 0.02) sets, respectively. Furthermore, in different age groups, this algorithm demonstrated consistently high performance, as suggested by median Dice score of 0.97 ± 0.02, 0.98 ± 0.02, 0.98 ± 0.01 in age categories of 0-4 years, 5-10 years, and 11-18 years.

**Table 1.** Performance metrics, including Dice score, sensitivity, and 95% Hausdorff distances on validation (val) set (n=43), withheld test (testi) set (n = 21), and independent test (teste) set from external sites (n = 29) by region: whole tumor (WT), enhancing tumor (ET), non-enhancing core (NEC) (i.e., a union of non-enhancing tumor, cystic component, and edema), and brain tissue.

| Region | Mean |  |  | Median |  |  | Standard Deviation |  |  |
| --- | --- | --- | --- | --- | --- | --- | --- | --- | --- |
|  | Val | Test<sub>i</sub> | Test<sub>e</sub> | Val | Test<sub>i</sub> | Test<sub>e</sub> | Val | Test<sub>i</sub> | Test<sub>e</sub> |
| **Dice Scores** |  |  |  |  |  |  |  |  |  |
| **WT** | 0.85 | 0.86 | 0.82 | 0.90 | 0.91 | 0.88 | 0.13 | 0.10 | 0.16 |
| **ET** | 0.67 | 0.68 | 0.74 | 0.75 | 0.73 | 0.84 | 0.30 | 0.27 | 0.29 |
| **NEC** | 0.74 | 0.73 | 0.62 | 0.78 | 0.79 | 0.74 | 0.15 | 0.19 | 0.27 |
| **Brain Tissue** | 0.98 | 0.97 | 0.97 | 0.98 | 0.98 | 0.98 | 0.01 | 0.02 | 0.02 |
| **Sensitivity** |  |  |  |  |  |  |  |  |  |
| **WT** | 0.85 | 0.84 | 0.83 | 0.90 | 0.89 | 0.89 | 0.14 | 0.13 | 0.18 |
| **ET** | 0.72 | 0.66 | 0.85 | 0.84 | 0.80 | 0.91 | 0.23 | 0.29 | 0.14 |
| **NEC** | 0.75 | 0.71 | 0.64 | 0.76 | 0.79 | 0.72 | 0.16 | 0.24 | 0.25 |
| **Brain Tissue** | 0.99 | 0.98 | 0.97 | 0.99 | 0.98 | 0.98 | 0.01 | 0.02 | 0.03 |
| **95% Hausdorff Distance** |  |  |  |  |  |  |  |  |  |
| **WT** | 9.51 | 10.08 | 10.93 | 3.74 | 3.74 | 3.61 | 16.47 | 13.65 | 14.07 |
| **ET** | 8.15 | 8.64 | 7.49 | 3.00 | 2.92 | 3.00 | 12.37 | 11.32 | 16.11 |
| **NEC** | 8.58 | 6.91 | 12.99 | 3.74 | 4.00 | 5.22 | 12.44 | 10.53 | 15.84 |
| **Brain Tissue** | 3.01 | 3.31 | 5.09 | 2.34 | 2.91 | 3.61 | 1.53 | 1.90 | 5.14 |

We further explored the advantage of applying a pediatric-specific model, as proposed in this study, compared to a model trained on adult patients on a few select patients in our cohort from different age groups. For this purpose, an adult brain extraction tool developed using DeepMedic architecture on adult gliomas, which is made available as an open-access and widely used module in CaPTk, was applied on the select patients. Figure S1 (Supplementary Material) on four patients, two in age range of 0-4 years, and two in the age range of 11-18 years suggests that applying an adult brain extraction model to pediatric brain scans may lead to under- or over-segmentation of brain tissue in certain regions.

### Tumor Subregion Segmentation

Performance metrics, i.e., Dice, sensitivity, and 95% Hausdorff Distance, on the validation, and internal and external test sets for the whole tumor (defined as the union of all segmented subregions), enhancing tumor, and the combination of all remaining subregions other than enhancing tumor, are summarized in Table 1. Figure S2 in Supplementary Materials demonstrates examples of predicted tumor subregion segmentation in two patients with low-grade glioma from the withheld internal test and external test cohorts compared to the expert segmentation (as the ground truth). Furthermore, this figure includes a boxplot of the Dice scores on the validation, and internal and external test sets for whole tumor, enhancing tumor, and non-enhancing core regions.

### Whole Tumor Segmentation

The mean Dice score of 0.85 on the whole tumor segmentation of the validation set, 0.86 on the internal test set, and 0.82 on the external test sets indicate strong agreement between the predictions obtained by our proposed model and the ground truth segmentations. For all subregions as well as whole tumor, the scores on the withheld internal test set were very close to those on the validation set, which positively indicates reproducibility of the model and its potential to perform well on unseen data.

As shown in Table 1, the median Dice scores of each tumor region on all sets were higher than the mean (0.90, 0.91, and 0.88 for the validation, and internal and external test sets, respectively). This result indicates that the model performs well on most patients but poorly on a few. On patients with lower Dice scores, mostly under-segmentation of the tumor was noticed.

The low 95% Hausdorff distances for the whole tumor (median of 3.74 on the validation set, and 3.74 and 3.61 on the internal and external test sets, respectively) suggest that the predicted boundaries were not far off from the ground truth boundary. In practice, these results suggest that most patients would require little to no manual revision of the whole tumor boundary, but a few may need and accurate whole tumor volume measurement, but some revision of subregions.

### Enhancing Tumor Segmentation

As indicated by the results in Table 1, for enhancing tumor, the model returned mean Dice of 0.67 on validation, 0.68 on the internal test, and 0.74 on the external test sets. Model performance on the enhancing tumor on the external test set was slightly better than validation and internal test sets, indicating generalizability of the model to the data from other centers. As observed with whole tumor, the median Dice scores were higher than the means, at 0.75, 0.73, 0.84 for the validation, internal test and external sets.

### Non-Enhancing Core

The average Dice scores on the combination of the three non-enhancing regions, i.e., non-enhancing tumor (NET), cystic component (CC), and peritumoral edema (ED), were 0.74, 0.73, and 0.62 on the validation, internal test, and external test cohorts, and higher than that of each of the three subregions individually. Mean Dice score (0.62) was lower on the external data, while median Dice (0.74) was close to the validation and internal test data (0.78, and 0.79, respectively). For patients where the model correctly identified a voxel as one of these three subregions but predicted the wrong label, sometimes non-enhancing tumor was mislabeled as a cyst (or vice versa) and sometimes non-enhancing tumor was mislabeled as edema (or vice versa). On the validation set, the mean ± standard deviation of Dice scores were 0.51 ± 0.30 for NET, 0.49 ± 0.34 for CC, and 0.31 ± 0.40 for ED. On the internal/external test sets, the scores were 0.51 ± 0.32 / 0.45 ± 0.27 for NET, 0.49 ± 0.40 / 0.46 ± 0.36 for CC, and 0.35 ± 0.43 / 0.31 ± 0.41 for ED. Of all subregions, the model exhibited lowest sensitivities and highest 95% Hausdorff distances on edema.

Figure 3 demonstrates two example cases in the withheld internal test set, for which the model was challenged in differentiating peritumoral edema and non-enhancing tumor from each other while showing accurate performance for segmentation of the whole tumor (Dice ≥ 0.90), enhancing tumor (Dice ≥ 0.83), and non-enhancing core (union of non-enhancing tumor, peritumoral edema, and cystic component) (Dice ≥ 0.83). In panel A, a small amount of the non-enhancing tumor was labeled as edema by the model, and in panel B, the near entirety of edema was labeled as non-enhancing tumor by the model.

**Figure 3.**
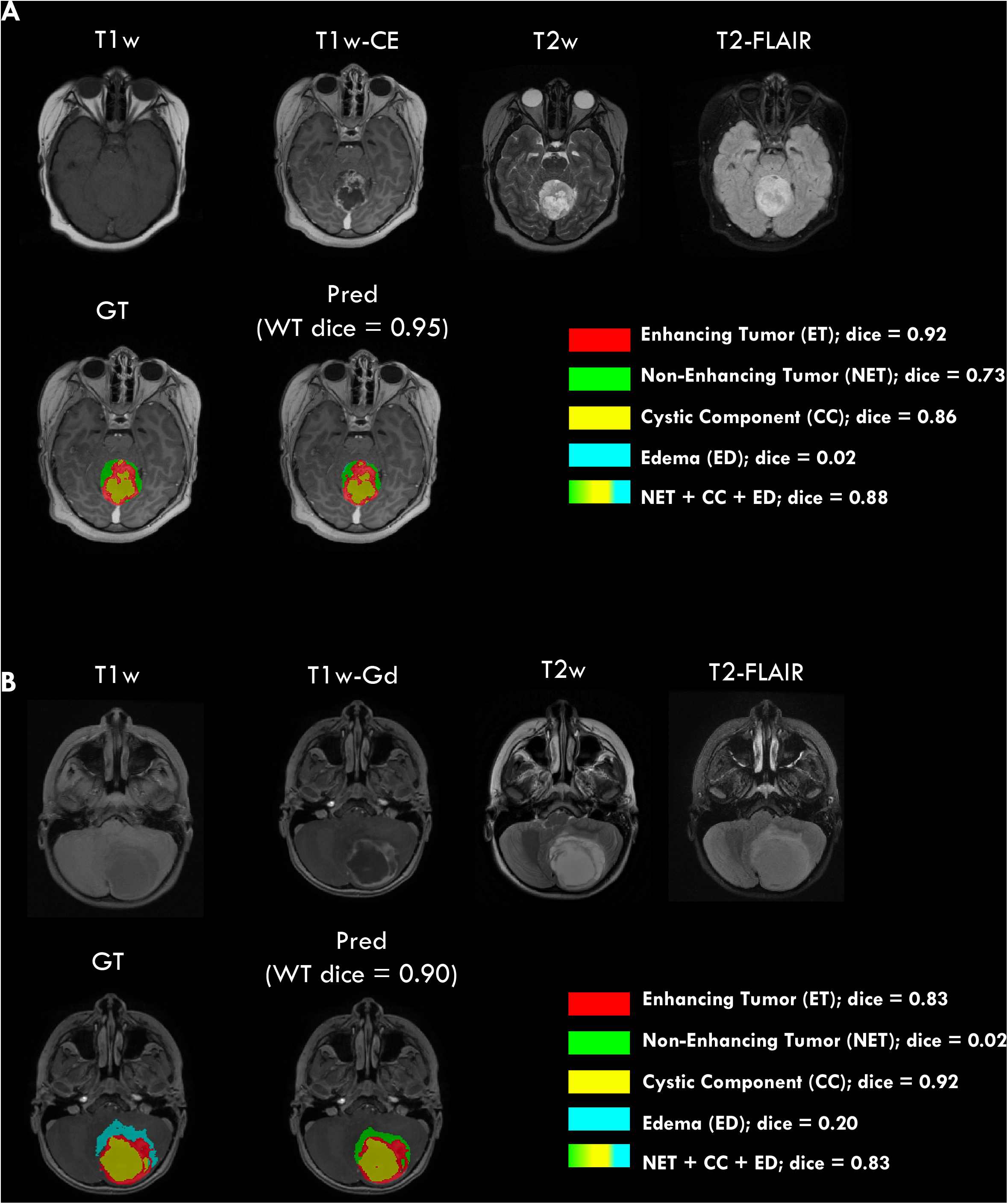
Examples of two subjects with accurate whole tumor, enhancing tumor, and non-enhancing core (combination of non-enhancing tumor, edema, and cystic component) but with low Dice scores on the non-enhancing tumor and edema subregions.

For the first patient in panel A, the predicted and manual segmentations showed excellent agreement of the whole tumor region, excellent sub-label identification of enhancing tumor and cyst, and good identification of non-enhancing tumor. The near-zero Dice score of edema and slightly lower non-enhancing tumor Dice score are caused by the model predicting a small amount of edema instead of non-enhancing tumor. For the second patient in panel B, the predicted and manual segmentations again showed excellent agreement of the whole tumor region as well as excellent sub-label identification of enhancing tumor and cystic components. The near-zero Dice scores of non-enhancing tumor and edema are due to the model predicting non-enhancing for nearly the entirety of the edema tumor region.

### Agreement of Model Predictions with Ground Truth for Tumor Volume

Further evaluation of agreement between model predictions (automated) and ground truth (manual) segmentations was carried out by calculating the ratio of the volume of the subregions to the whole tumor region for the patients in validation, internal test, and external test cohorts (Figure 4). The correlation coefficients were R = 0.94 (p<0.01) for ET/WT, R = 0.86 (p<0.01) for NET/WT, R = 0.8 (p<0.01) for CC/WT, and R = 0.47 (p<0.05) for ED/WT. Agreement between the automated and manual segmentations for measuring these volume ratios are summarized in Bland-Altman plots in Figure 5. The mean difference and 95% limits of agreement (summarized as mean difference [95% lower limit – 95% upper limit]) between automated and manual segmentation methods for the proportion of each subregion volume to whole tumor volume were as follows: ET/WT, 0.01 [-0.19 – 0.22]; NET/WT, 0.01 [-0.35 – 0.38]; CC/WT, 0.00 [-0.20 – 0.20]; ED/WT, -0.02 [-0.28 – 0.23].

**Figure 4.**
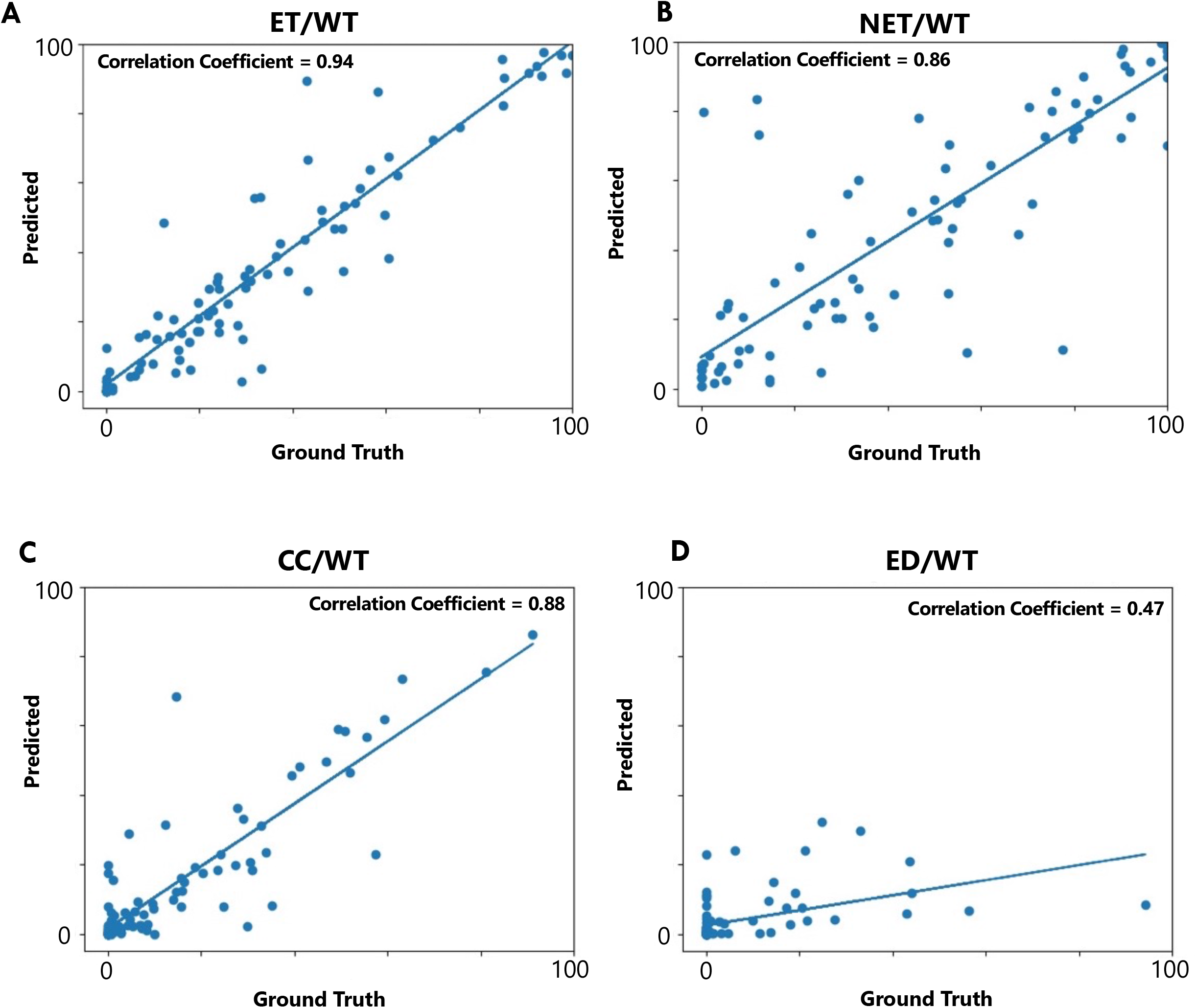
Scatter plot of results from comparison of the agreement between the volumes of the tumor subregions predicted by our proposed tumor subregion segmentation algorithm compared to those from the ground truth: (A) VASARI feature f5, i.e., proportion of tumor that is enhancing; (B) Vasari feature f6, i.e., proportion of tumor that is non-enhancing; (C) the proportion of tumor that is cystic, which is not a VASARI feature. (D) VASARI feature f14, i.e., proportion of tumor that is edema.

**Figure 5.**
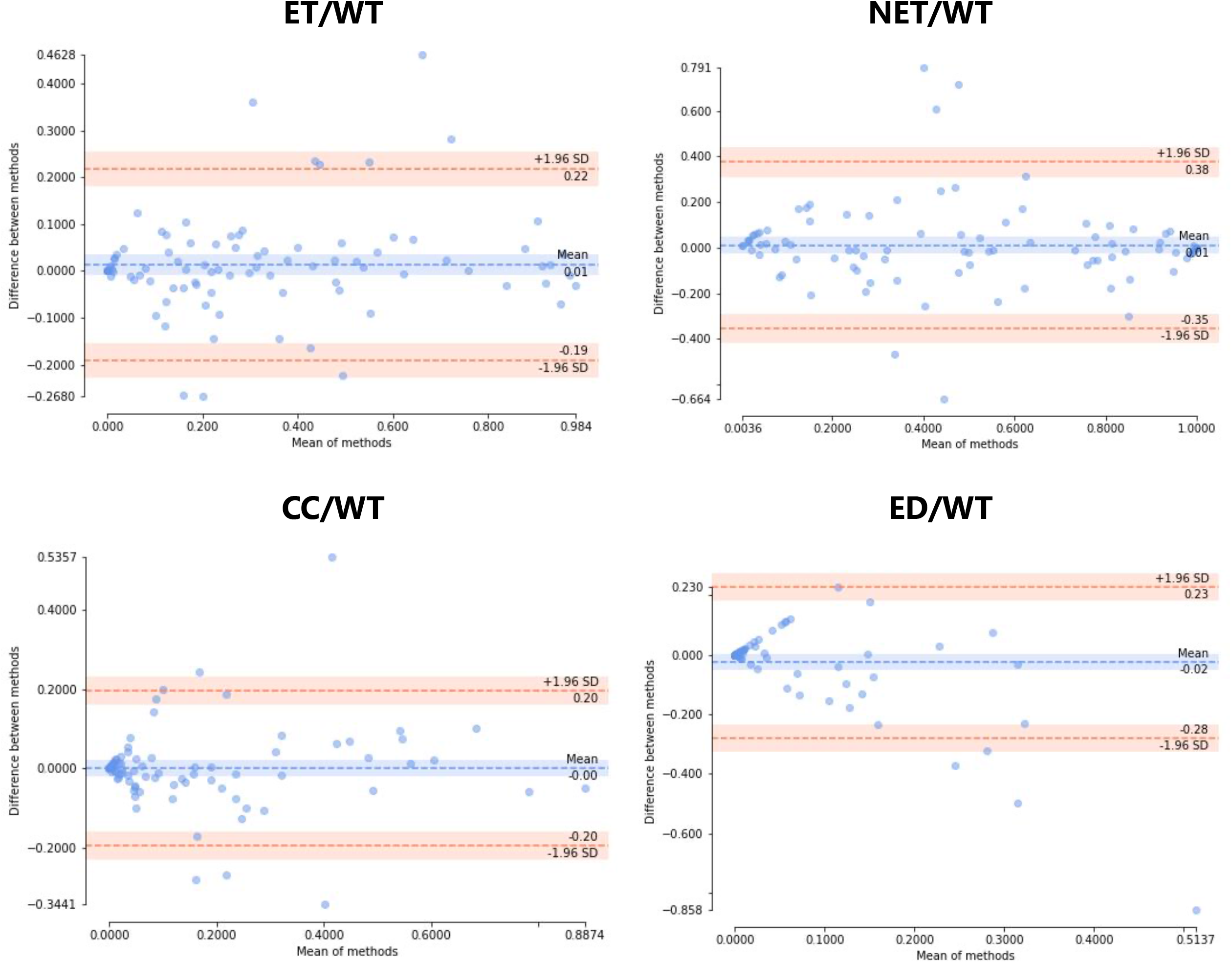
Bland-Altman plots illustrating agreement of our proposed segmentation method with manual segmentation in measuring the ratio of volume of each tumor subregion to volume of whole tumor for the patients in internal validation and test, and external test cohorts.

## Discussion

Accurate whole tumor segmentation and reliable subregion differentiation is critical for volumetric measurements for surveillance of tumor progression and response assessment ^6^, as well as for radiomic studies in neuro-oncology ^25^. However, the process of manually generating 3D segmentations imposes high manual burden on the readers and is prone to variability and subjectivity ^8^. In this study, we presented multiparametric, multi-scanner, multi-histology, and multi-institutional deep learning-based models for brain tissue extraction and tumor subregion segmentation, that to the best of our knowledge, are the first deep learning methods to automatically skull-strip and segment pediatric brain tumor subregions.

The proposed models in this study yielded excellent brain tissue extraction and whole tumor segmentation performances, as well as decent performance on enhancing and non-enhancing tumor subregions. The results suggest that this model could be used to provide a useful preliminary segmentation that would largely reduce the time burden on the radiologist by only requiring simple revisions. The proposed brain extraction model achieved Dice score of ≥0.97 on all data subsets, confirming its reproducible and generalizable performance to the unseen data from internal and external cohorts. Availability of a pediatric-specific model is preferred over applying existing the pre-trained models generated based on adult brain tumors, as the structure and MR image signal intensities vary largely in pediatric cohorts with developing brains ^16^.

In the present study, the tumor segmentation model produced median Dice scores of 0.90 on the validation, 0.91 on the internal test, and 0.88 on the external test sets for delineation of whole tumor region. These results are significantly higher than other pediatric deep learning models ^11–13^, which achieved Dice scores ranging from 0.71 to 0.76. Several of these existing models are focused on a particular histology, such as optic pathway gliomas (OPGs) ^17,20^. The variety of histologies and tumor locations included in our model allowed us to build a model that can be used more generally. This may be specifically useful if the tumor diagnosis, in a prospective patient, is not yet known or if a dedicated model for a particular histology does not exist, as is typically the case in clinical contexts. In other words, it could provide an accurate whole tumor segmentation on an unseen patient, while a model dedicatedly trained on a more homogenous cohort (e.g., all OPGs) may not generalize well to the images of a patient that differs too much from its training set (e.g., a high-grade glioma or a tumor located outside the optic system). Additionally, our large patient cohort size positively contributed to the ability of the model to perform well on unseen data.

As RAPNO guidelines are more widely being adopted for clinical research, it will become more essential to segment tumors according to these guidelines. An automated segmentation approach for PBT subregion segmentation has the potential to reduce variability across manual segmentations and help standardize segmentations across radiologists and institutions in clinical trials. In this light, our proposed automated deep learning-based tumor segmentation model can be further used to provide measurements of the tumor (subregion) volume for clinical assessment, i.e., tracking tumor progression per RAPNO guidelines. This model showed strong agreement with manual segmentation in estimating the volumetric ratios of tumor subregions to the whole tumor volume. This was confirmed by the strong correlation obtained for the volume ratios of ET/WT, NET/WT, and CC/WT, and moderate correlation for ED/WT between the automated and manual segmentations. Some of these features representing volumetric ratios are among well-known VASARI features, and are commonly used as semantic radiomic features reported in previous studies ^26^.

Bland-Altman plots, to evaluate agreement between the auto-segmentation and manual segmentation methods for measuring ET/WT, NET/WT, CC/WT, and ED/WT volume ratios, further indicated little bias in volumetric measurements using our proposed automated segmentation method, as suggested by near-zero mean difference in all volumetric ratios. For only a few subjects the measurements of the automated segmentation method fall outside of the limits of agreement. Altogether, these results suggest that the automated segmentation method can be used reliably for tumor measurements, as a substitute for a fully manual segmentation approach. Manual revision may still be needed in some cases to refine whole tumor and component boundaries, and differentiate between cystic core and cystic reactive components, if necessary. Nonetheless, the manual burden will be considerably reduced when using the segmentations provided by such automated segmentation methods. A downstream benefit of such automated brain tissue and tumor segmentation tools is to provide an automated tool for radiomic analysis studies, that could facilitate better replication of the generated radiomic models to data from different institutions through standardization of segmentations and therefore the resulting features.

The model performed well on segmentation of enhancing tumor (median Dice of 0.75, 0.73, 0.84 on the validation, internal test, and external test sets, respectively) and non-enhancing core (median Dice of 0.78, 0.79, 0.74 on the validation, internal test, and external test sets, respectively). However, in some subjects, the assignment of pixels to the enhancing and non-enhancing core regions was suboptimal. On patients with lower Dice scores on enhancing tumor, a few trends were noticed. First, the model may have predicted just a few voxels of enhancing tumor, and the ground truth segmentation had none, which results in a calculated Dice score of zero. Second, the ground truth segmentation may have had an enhancing region that was very mildly enhancing, and the prediction model did not label the region as enhancing. This challenge in model performance occurred particularly when baseline differences existed in relative signal intensities of normal brain tissue on pre-contrast compared to post-contrast T1w scans. Another challenging factor in this regard was subjectivity in labeling the voxels with slight enhancement and variability in their assignment to the enhancing tumor or non-enhancing core. In general, with a few exceptions, the errors in prediction of the model on enhancing tumor were minor and would require only minor manual revision. The high sensitivity and low 95% Hausdorff distances also support this finding. While the model provides a decent preliminary tumor subregion differentiation to provide far less time-consuming manual revisions on the predicted segmentations than a segmentation from scratch by easily changing the labels to correct the subregion assignment, the model performed poorly in segmentation of some of the non-enhancing tumor components from each other. Inspection of such cases indicated that the automated DL model may not perform well on cases which expert readers may also find difficult to determine. For instance, one factor that may have negatively influenced the performance of the model is that manual differentiation between edema and non-enhancing tumor is challenging, as there is substantial overlap in the radiological appearance of edema and non-enhancing tumor on standard (or conventional) MRI scans. Similarly, if a cystic component does not suppress heavily on T2-FLAIR, manual segmentation can sometimes be subjective in deciding between a cyst and non-enhancing tumor. Poor performance of the model on edema may also be partially explained when in manual segmentations, transependymal edema signal adjacent to ventricles due to hydrocephalus was purposefully not segmented as tumor edema, causing issues for the algorithm learning. Thus, model would have to learn to differentiate peritumoral edema based on anatomical location along with imaging appearance, which is typically utilized by convolutional networks such as the one employed here. Another likely contributing factor to this result was class imbalance, for example, edema was only present in 44.6% of the tumors, which limits the examples that the model can learn from.

Furthermore, the Dice score, as calculated here, is not an ideal metric for understanding the performance of the model on rarer labels (i.e., labels only present in a subset of the samples). For instance, if the predicted segmentation has 10 voxels of edema, and the ground truth does not have any edema, that will return a Dice score of zero. On the other hand, if the ground truth had 20,000 voxels of edema, and the model predicted no edema, that will also return a Dice score of zero. The latter theoretical situation is clearly worse because it would require more manual revision than the former, especially when the whole tumor volume is often on the scale of tens of thousands of voxels. The reverse is also true – a Dice score of 1 could indicate 10 out of 10 voxels segmented correctly, or 30,000 out of 30,000. Ideally, performance metrics would indicate the accuracy of the segmentation in a way that conveys the severity of the mistake and the importance of a correct prediction. In our study, we chose the standard method of calculating dice score to compare with other studies on automatic brain tumor segmentation.

While one of the benefits of this work was inclusion of multi-institutional and multi-scanner data, variabilities in scan protocols may have contributed to suboptimal performance in segmentation of some of the regions. To mitigate the effects of this variability, we applied a standardized image processing approach to align the images and normalized the image intensities; however, variance in scanner acquisition parameters can nonetheless influence image quality. Increasing the cohort size could further help in minimizing the influence of such variabilities and improving the model performance in tumor subregion segmentation to the point where segmentation editing is minimized or no longer needed in most cases.

In conclusion, we presented automated deep learning-based brain extraction and tumor subregion segmentation models based on a multi-institutional dataset. The developed models can reliably segment the MRI scans of the children across a variety of brain tumor histologies and based on widely available standard clinically acquired MRI scans. Such non-invasive methods can offer rapid delineation of tumor from surrounding regions and provide measurement of tumor volume with more consistency, and less inter-observer variability than manual segmentation methods. As a result, the automated DL model can be applied across clinical and research settings to provide standard tumor measurements and correspondingly monitor tumor progression and treatment response.

## Data Availability

All data produced in the present study are available upon reasonable request to the authors

