## Supplemental Table and Figures for "Automated Tumor Segmentation and Brain Tissue Extraction from Multiparametric MRI of Pediatric Brain Tumors: A Multi-Institutional Study"

**Table S1.** Patient characteristics in the studied cohorts (internal cohort of 215 and external cohorts of 29 patients)

| **Patient Characteristics** | **Internal Cohort** | **External Cohorts** |
| --- | --- | --- |
| Total Patients | 215 | 29 |
| Age at imaging, range (years) | 0.24 to 20.15 | 0.53 to 19.70 |
| Age at imaging, median (years) | 7.76 | 9.35 |
| Gender | 113 Male, 102 Female | 13 Male, 16 Female |
| Histology |  |  |
| Low-Grade Glioma / astrocytoma | 119 | 21 |
| High-Grade Glioma / astrocytoma | 21 | 3 |
| High-Grade Glioma / DMG (pontine location) | 20 | 0 |
| Ependymoma | 5 | 0 |
| Medulloblastoma | 47 | 4 |
| Central neurocytoma | 0 | 1 |
| Histiocytic Neoplasm with BRAF V600E mutation  Germinoma | 1  2 | 0  0 |
| Scanner Magnetic Field Strength (T) |  |  |
| 0.7  1.5  3  Information unavailable  Scanner Manufacturer  Siemens  GE  Phillips  Toshiba | 0  57  154  4  200  13  0  1 | 1  19  9  0  11  13  5  0 |
| Hitachi | 1 | 0 |

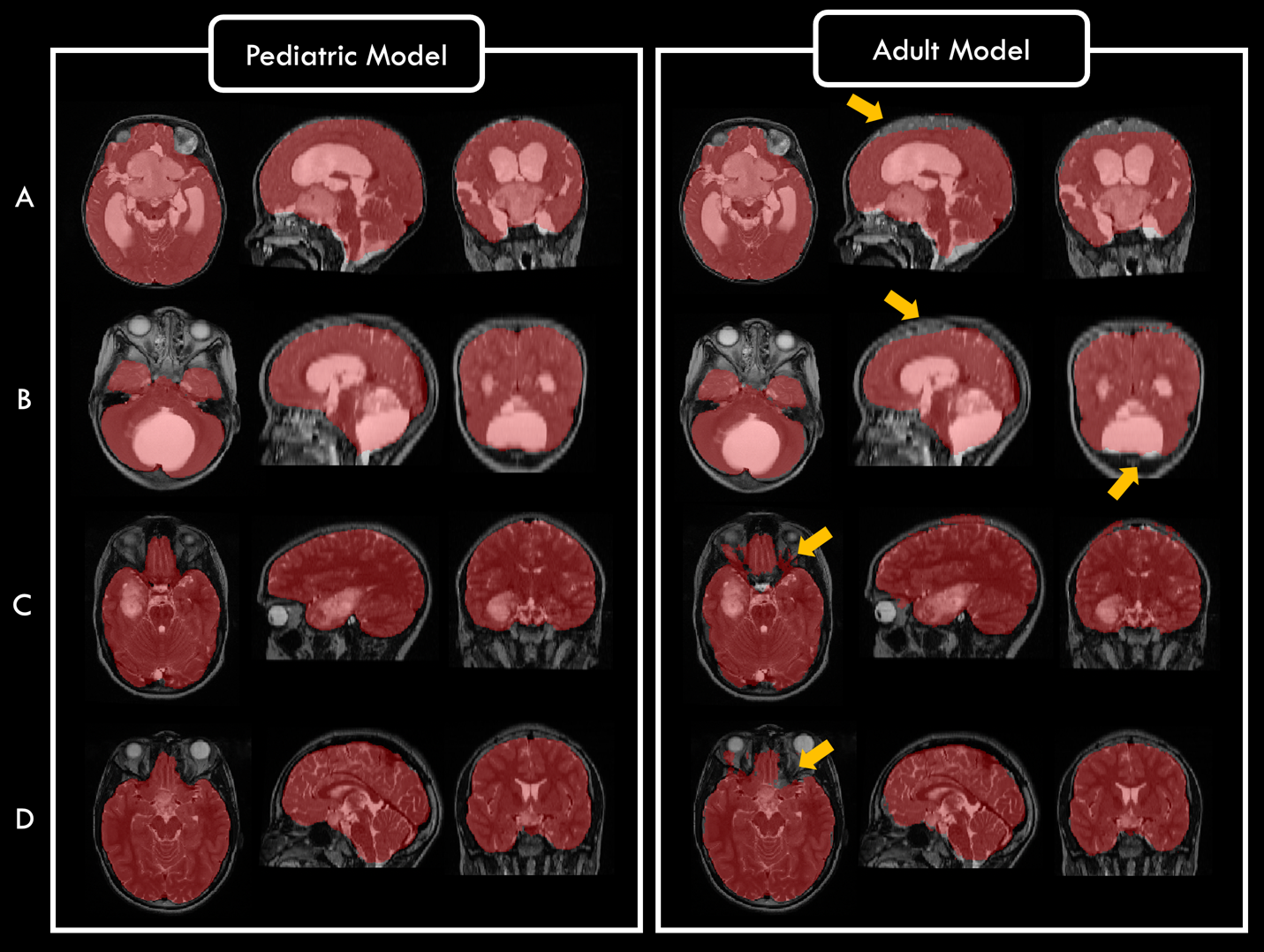

**Figure S1.** Brain tissue extraction using our proposed pediatric DeepMedic model, developed on pediatric brain tumors, compared to an adult DeepMedic model trained on adult gliomas. Each panel (pediatric or adult model) shows T2w images in the axial, coronal, and sagittal planes with the predicted brain masks overlaid. Yellow arrows on the right panel (adult model) point to the under or over-segmented regions. The scans are selected from T2w images of two patients in age range of 0-4 years (panels A & B), and two patients in age range of 11-18 years (panels C & D).

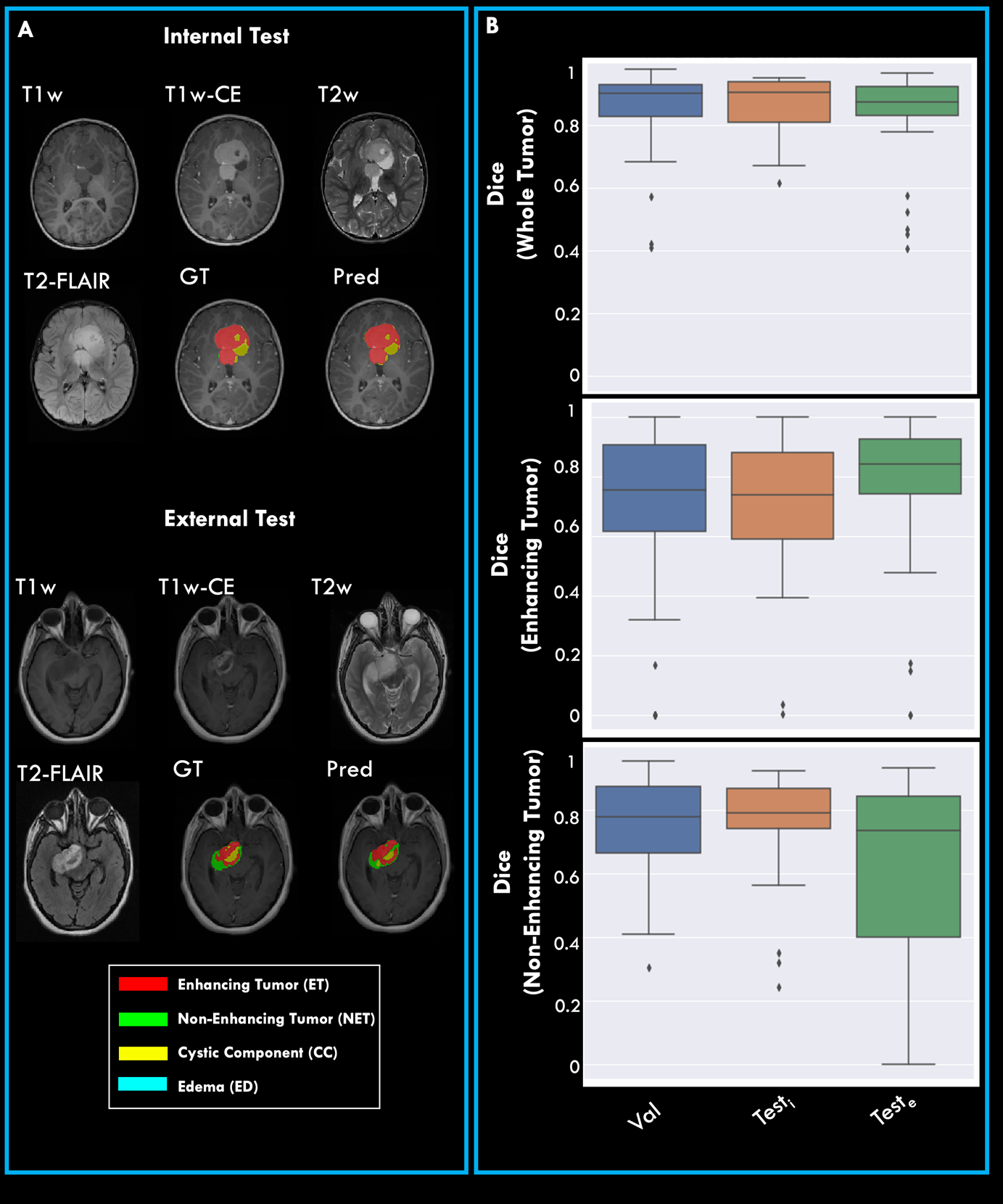

**Figure S2.** Tumor subregion segmentation: Panel (A) demonstrates multiparametric scans along with predicted (Pred) tumor subregions using our proposed algorithm and expert segmentation (ground truth, GT) overlaid on T1w-CE scan for two patients with low-grade glioma. The scans are for a patient from the withheld test set and the scans at the bottom belong to another patient from the external test cohorts. Panel (B) shows boxplots for the Dice scores computed for predicted tumor subregion segmentation in validation (Val), internal test (Test_i_) and external test (Test_e_) subjects for whole tumor, enhancing tumor, and non-enhancing core regions.
